# A Device to Prevent Night-time Sudden Unexpected Death in Epilepsy

**DOI:** 10.1101/2024.01.02.23300653

**Authors:** Jong Woo Lee, Pranav Bansal, Ioannis Smanis, Justin Nguyen, Andres Rodriguez

## Abstract

**Background:** Sudden Unexpected Death in Epilepsy (SUDEP) is the leading cause of death in epilepsy children and otherwise healthy adult epilepsy patients. About 70% of SUDEP occurs during sleep, and nearly 90% are found in the prone (face-down) position. SUDEP can likely be prevented by simple interventions such as turning and stimulating. Such intervention must be performed quickly within a 3-minute window prior to death. There are currently no products that detect the prone position or have the ability to physically reposition a patient into a recovery position.

**Methods:** We address this critical unmet medical need with a smart mattress, the Korus, consisting of an array of inflatable cells. We developed 1) a sensor system to rapidly detect body change with high accuracy; 2) an advanced expandable cell that is capable of generating the required lift at sufficient velocity to reposition a patient from the prone to the recovery (sideways) position.

**Results:** Each inflatable cell that comprises the Korus has the capacity to generate lift to 1000 lb/cell at a rate of 2 inches per second. The embedded sensor array can detect body position change within 5 seconds. A total of 10 normative control subjects were tested. Repositioning from a prone to recovery (sideways) position while manually controlling the cells was successful in 100% of attempts with an average time of 21.75 sec. Body position detection resulted in an accuracy in detecting the prone position of 96.8% and an overall accuracy of detecting the correct body position (supine, prone, left, right) of 92.4%. In long-term studies including sleep, Korus detected the correct position during 97.3% of the time during the recording. False body position change rate was 2.3 events/hour. There were no incorrect position detections when subjects switched into the prone position.

**Discussion:** This study demonstrates the feasibility of developing a smart mattress to detect the prone position and rapidly repositioning subjects into a recovery position. Future development include the development of a control system to automate repositioning based on sensor data. The completion of this device, when paired with a seizure detection device, has the potential to lower the risk of SUDEP by >50%.

## I. Introduction

Sudden Unexpected Death in Epilepsy (SUDEP) is the non-traumatic and non-drowning death of a person with epilepsy. It is the leading killer of healthy young patients with epilepsy and the second-leading neurological cause of total years of potential life lost after stroke.(1) The risk of SUDEP is highest in the ∼one million patients whose seizures are not controlled by medication(2), but patients with benign epilepsy conditions are also at risk.(3) Although 20 new antiseizure medications have been introduced over the past 30 years, the probability of achieving seizure freedom has not changed.(4) There is an under-diagnosis(5) and an increasing awareness and fear of SUDEP.(6) There is a clear need for an intervention to reduce the risk of SUDEP.

The strongest risk factor for SUDEP is poor control of generalized tonic-clonic seizures (GTCS).(2) A GTCS is followed by a depressed level of consciousness or impaired arousal. Peri-ictal respiratory dysfunction is characterized by a combination of poor respiratory mechanics, arousal failure, and decreased respiratory drive, leading to apnea, or cessation of respiration, within approximately 3 minutes.(7)

### Convulsive seizures during sleep and prone position are major risk factors for SUDEP

A meta-analysis of 880 patients with SUDEP revealed that ∼70% of SUDEP occurs after a nocturnal generalized tonic-clonic seizure (GTCS)(8–10) due to greater cardiorespiratory instability during sleep, postictal airway obstruction, and increased likelihood of being alone.(11) Up to 87.6% of patients are found in the prone (face-down) position after a nocturnal SUDEP(8,12), despite the fact that the incidence of prone position in the post-ictal state of GTCs is an uncommon event, occurring at rates of only 4.2-7.2%.(13,14) Central apnea(10,15,16) and autonomic instability(17–19) have been identified as key risk factors for SUDEP. Prone sleeping position blunts arousal responses and reduces heart rate variability.(20–22) Through these factors, the prone position potentially contributes to both central apnea and autonomic instability. The relative risk of SUDEP in the prone vs non-prone position following a GTCS is over 60-fold.(23)

### Convulsive seizures during sleep and prone position are major risk factors for SUDEP

A meta-analysis of 880 patients with SUDEP revealed that ∼70% of SUDEP occurs after a nocturnal generalized tonic-clonic seizure (GTCS)(8–10) due to greater cardiorespiratory instability during sleep, postictal airway obstruction, and increased likelihood of being alone.(11) Up to 87.6% of patients are found in the prone (face-down) position after a nocturnal SUDEP(8,12), despite the fact that the incidence of prone position in the post-ictal state of GTCs is an uncommon event, occurring at rates of only 4.2-7.2%.(13,14) Central apnea(10,15,16) and autonomic instability(17–19) have been identified as key risk factors for SUDEP. Prone sleeping position blunts arousal responses and reduces heart rate variability, potentially contributing to both.(20–22) The relative risk of SUDEP in the prone vs non-prone position following a GTCS is over 60-fold.(23)

### Convulsive seizures during sleep and prone position are major risk factors for SUDEP

A meta-analysis of 880 patients with SUDEP revealed that ∼70% of SUDEP occurs after a nocturnal generalized tonic-clonic seizure (GTCS)(8–10) due to greater cardiorespiratory instability during sleep, postictal airway obstruction, and increased likelihood of being alone.(11) Up to 87.6% of patients are found in the prone (face-down) position after a nocturnal SUDEP(8,12), despite the fact that the incidence of prone position in the post-ictal state of GTCs is an uncommon event, occurring at rates of only 4.2-7.2%.(13,14) Central apnea(10,15,16) and autonomic instability(17–19) have been identified as key risk factors for SUDEP. Prone sleeping position blunts arousal responses and reduces heart rate variability.(20–22) Through these factors, the prone position potentially contributes to both central apnea and autonomic instability. The relative risk of SUDEP in the prone vs non-prone position following a GTCS is over 60-fold.(23)

### SUDEP can be prevented by self-repositioning the patient

Patients can be saved from SUDEP by repositioning to avoid the postictal prone position: **1)** Sudden Infant Death Syndrome (SIDS), the closest analogous disorder to SUDEP sharing many pathophysiologic features(7,24,25) and similar risk with prone position(26,27); has been reduced by up to 75% through “back to sleep” prevention campaigns.(26,28–30) **2)** Every SUDEP documented by video-EEG has died in the prone position.(10,31–33) Asphyxia from airway obstruction and hypoventilation due to a patient’s prone positioning has been documented on video-EEG.(31) **3)** At home, just having nocturnal supervision or a bed partner decreases the risk of SUDEP(34–37), suggesting that simple interventions such as repositioning and stimulating the patient substantially decrease risk.

### Timing of intervention is critical

In convulsions leading to SUDEP, terminal asystole and apnea occur as soon as 25 sec postictally and in virtually everyone by 180 sec(10). Intervention must occur during this time. One patient died of SUDEP wearing the Empatica seizure detection device (considered best in class) that was functioning flawlessly; he was found prone in bed despite help arriving in 15 minutes.(38) Autonomous intervention is therefore imperative. There are currently no methods to deliver interventions within the required time.

An effective intervention to prevent SUDEP, particularly after a nocturnal convulsion, is clearly needed as medications will not control seizures in a large portion of patients with epilepsy. The overarching goal of this study is to develop a night-time seizure management system centered around an autonomous body repositioning mattress after a GTCS. Here, we report results of the development of this device, Korus.

## II. Materials and Methods

### IRB Approval

This study was approved by the Mass General Brigham institutional review board under protocol 2021P002558.

### Description

The mattress of size of a twin XL bed consists of an array of inflatable cells that expand 4x, up to 14” in the Z axis. Cells are distribute based on matrix of an 8 by 4 cells with three main sections; pad section where several layers of foam form a supportive surface and where the sensor is located, the spring section, and the base section where cell controller and valves are located. Cells are placed on a base board that contains data, power, and air distribution lines or networking system. This arrangement constitutes the structure and body of the mattress and forms a shapable and sensing surface suitable for sleeping and repositioning; The cells can be detached from the baseboard to be replaced or fixed individually. The arrangement is wrapped in a mattress cover that stretches to allow the formation of the shapes along the sleeping surface. This cover uses several layers of fabric such as spandex, neoprene, and other cotton and anti-fluidics fabrics.

#### A. Inflatable cells

The cell was designed considering several factors including human factors such as diversity of patients’ body types, weight demands, and sleep experience requirements. Federal regulations have been considered including Flammability Standards: 16 CFR Parts 1632 and 1633, FDA recommendations for Mattresses and Pillows G.VII., and durable medical equipment (DMEPOS) quality and requirement standards. The core of the cell is an inflatable spring that expands up to 14”. The manufacturing processes involved injection and vulcanizing molding using natural rubber and nylon cord. These materials are widely used in the medical industry and offer good mechanical properties such as temperature-stability, no toxic or irritant, can be sterilized, chemical stability, resistance to hydrocarbons, oils and solvents, flexibility/elasticity, and moldability. Two components constitute the core of the expandable function of the cell. These components participate in the control and regulation and contention of the air allowing the expansion of the cell: A set of solenoid valves that regulate the air flow in and out of the spring.

The systems feature inflation and deflation for greater control over the shaping of the sleeping surface. A safety air-valve was included to prevent over inflation of the spring. The spring has a maximum working pressure of 100 Psi. However, the system is set to a max working pressure of 35 Psi. For manufacturing plastic parts, 3D printing was performed using several technologies including materials such as PLA and PETG filaments. Computer numeric cutting (CNC) was used for bigger parts such as the based board on wood and foam.

#### B. Sensors and networking system

The main sensor in the bed is a sheet sensor embedded into cell pad in each cell. The sheet sensor is ∼2 mm thick with flexible circuitry with 36 nodes per sensor which forms a pressure sensing surface of 1,152 nodes in the XL twin size bed with 256 pressure levels. The networking platform features a Power over Ethernet (PoE) and three main custom made electronic boards: Sensor sheet, sensor controller, and cell controller. Other components include PoE switch boxes, an internet router, and a Raspberry Pi 8GB for computing. The entire system interacts within a local network where each cell has an IP address and data packages include the pressure map and other data outputs from additional sensors in the cell such as accelerometers and temperature sensors.

#### C. Body position detection

The position recognition is performed using a Random Forest Classifier model which takes the whole set of pressure values as input and calculates a position based on them. Each node has 36 pressure values therefore there are 32*36 = 1152 values in the input vector for the model which helps the model accurately recognize the position. With the help of the training process, the model can save a map for these values for a specific user in different positions. The Random Forest model internally utilizes an ensemble of several decision trees. Each individual decision tree in the model generates a classification as output and the Random Forest Model combines all these outputs to generate a consolidated and more accurate result.

A machine-learning (ML) algorithm was developed to detect the 4 cardinal body positions (prone, supine, left, right).

#### D. App development

An iOS and Android app was developed to interact with the bed and visualize the body position. Body position is displayed in real time as an avatar.

However, to streamline the development process particularly the ML training process, a collection of data workflow was integrated into the app and an automatized training process was added to the back end. This facilitated and simplified the data gathering, training, and enhancement of the model by following a sequences of position changes and recording process. This process took less than 10 mins per subject. The app also allows the control of the smart cells; degree of inflation/deflation of each cell is individually addressable. The streaming of body position detection system, and a workflow.

#### E. Participants

A total of 18 normative control subjects were enrolled, of whom 10 (ages 18-53, weight 100-182, height 5’2”-6’1”) underwent extensive formal testing. This study was approved by the Mass General Brigham institutional review board.

## III. Results

### Cell performance

The cells achieved their target 14” inflation within 7 seconds (rate 2”/sec) with a 1000 lb/cell capacity. Recovery position in 10 normative control subjects from the prone position was achieved with a success rate of 100% with an average time of 21.75 +/-5.85 sec (range 15-29 sec) through manual control of the cells.

### Body position recognition

The overall accuracy in detecting the prone position over 10 patients was 96.8% [83.3-99.9]. There were no instances where supine and prone positions were misidentified for each other. Overall accuracy over all positions was 92.4% [86.1-96.5], with most of the errors arising erroneous detection of side positions (Figure 1).

**Figure 1.**
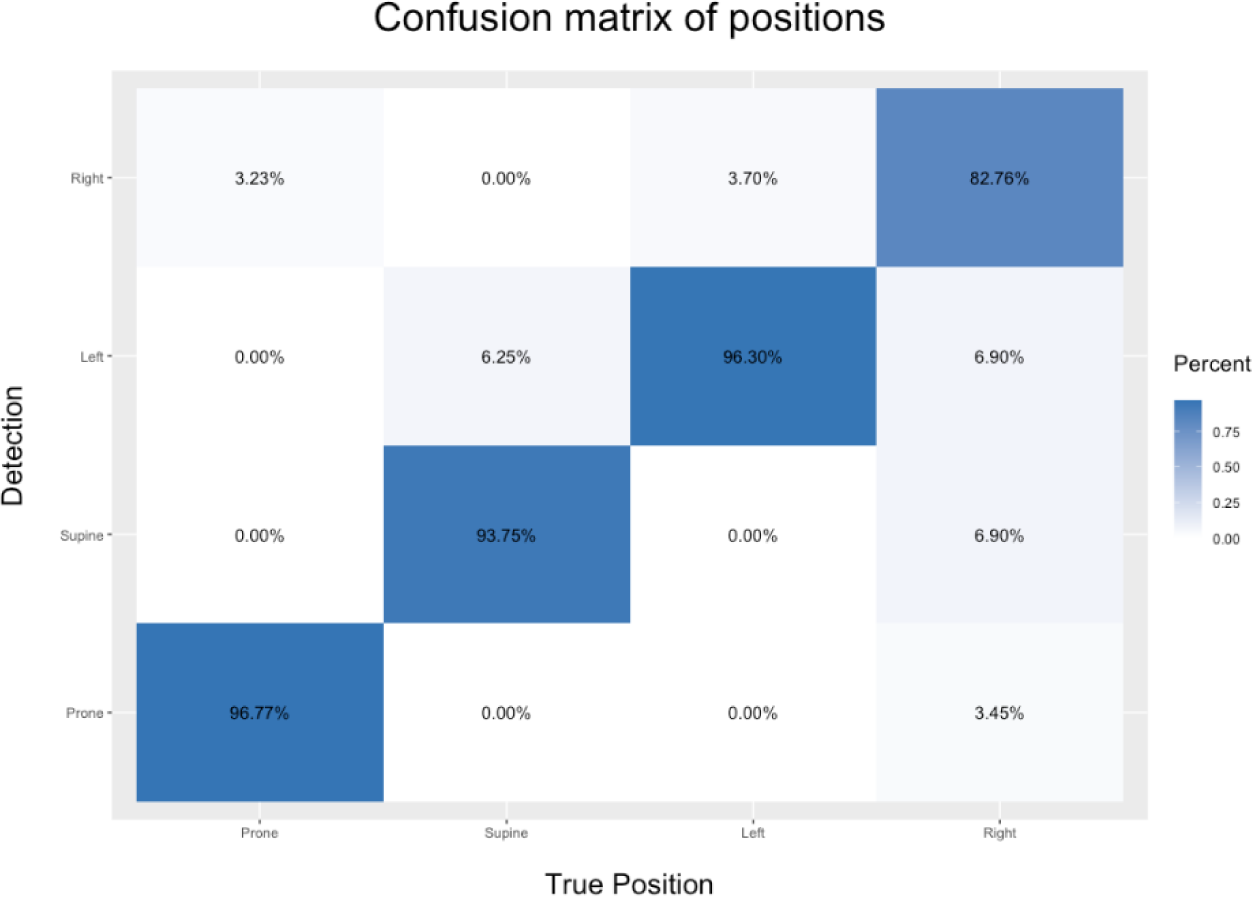
Confusion matrix demonstrating accuracy of the Korus body position detection sensors and algorithm. Detection of prone position had overall accuracy of 96.8%.

Inability to recognize the correct body position within 5 sec or without weight shifting (though maintaining the same orientation) were considered failures. Long-term recording with sleep revealed body position change at a rate of 1.67/hour [0.23, 2.14]. Korus detected the correct position during 97.3% of the time during the recording. False body position change rate was 2.3 events/hour [1.2, 4.1]; video review revealed this was always due to a supine position as being incorrectly recognized as left/right. There were no incorrect position detections into the prone position. Gross failure was noted in one subject due to poor body recognition due to the subject bending legs during sleep, which was not captured during the training session; this subject was not included in the analysis.

## IV. Discussion

An effective intervention to prevent SUDEP, particularly after a nocturnal convulsion, is clearly needed as medications will not control seizures in a large portion of patients with epilepsy. Although this need has been widely recognized, current available options remain insufficient. Seizure detection devices and remote listening strategies do not have the ability to intervene and prevent SUDEP on their own and will not allow for consistent delivery of rapid treatment within the critical window (**Table**). Daily use of such devices is challenging, given the need to remember to wear the device, charge the battery, and establish a reliable network for alerts. Anti-asphyxia pillow/mattress toppers have been developed to reduce airflow resistance and prevent suffocation, but carbon dioxide detention still exceeds 8%, which is considered potentially life-threatening.(39) The Korus has a first-in-class potential to monitor and autonomously deliver intervention to prevent SUDEP.

The current Korus model achieves both the necessary body repositioning task and recognition of the cardinal body position (prone, supine, left lateral, right lateral). Several tasks remain before a fully autonomous SUDEP prevention system is realized including the improvement of the quality of the map, optimization of the computing processes, and implementation of a biomechanical framework. The Korus can be paired with peripheral devices such as wearables for seizure detection. In addition, our development plan included the development of seizure detection algorithms with Korus sensing system. Realization of these goals would potentially achieve an expected effect size of SUDEP risk reduction of >50%, comparable to the risk reduction achieved by *strict* avoidance of prone positioning in SIDS.(40)

**Table.** SUDEP prevention devices and strategies

**Table. SUDEP prevention devices and strategies**
| Solutions | Widely available | Autonomous | Rapid intervention | No wearable required | Multiple treatments | Effectiveness |
| --- | --- | --- | --- | --- | --- | --- |
| Soterya's KORUS | ✓ | ✓ | ✓ | ✓ | Repositioning and stimulation | Up to 75% |
| Wrist-worn seizure detection device (Embrace) | ✓ | X | X | X | X | Unlikely by itself |
| Under-mattress device (MM) | ✓ | X | X | ✓ | X | Unclear |
| Breathable pillow (Sleep Safe) | ✓ | ✓ | ✓ | ✓ | X | Unclear |
| Central listening device<br>system | X | X | ✓ | ✓ | X | Est. 2.8-fold risk<br>reduction |

## Data Availability

All data produced in the present study are available upon reasonable request to the authors

## References

1. Thurman DJ, Logroscino G, Beghi E, Hauser WA, Hesdorffer DC, Newton CR, et al. The burden of premature mortality of epilepsy in high-income countries: A systematic review from the Mortality Task Force of the International League Against Epilepsy. Epilepsia. 2017;58:17–26. DOI: 10.1111/epi.13604.

2. Devinsky O, Hesdorffer DC, Thurman DJ, Lhatoo S, Richerson G. Sudden unexpected death in epilepsy: epidemiology, mechanisms, and prevention. The Lancet Neurology. 2016;15:1075–88. DOI: 10.1016/S1474-4422(16)30158-2.

3. Doumlele K, Friedman D, Buchhalter J, Donner EJ, Louik J, Devinsky O. Sudden Unexpected Death in Epilepsy Among Patients With Benign Childhood Epilepsy With Centrotemporal Spikes. JAMA Neurol. 2017/04/07 ed. 2017 Jun 1;74(6):645–9. DOI: 10.1001/jamaneurol.2016.6126.

4. Chen Z, Brodie MJ, Liew D, Kwan P. Treatment Outcomes in Patients With Newly Diagnosed Epilepsy Treated With Established and New Antiepileptic Drugs: A 30-Year Longitudinal Cohort Study. JAMA Neurol. 2017/12/28 ed. 2018 Mar 1;75(3):279–86. DOI: 10.1001/jamaneurol.2017.3949.

5. Devinsky O, Friedman D, Cheng JY, Moffatt E, Kim A. Underestimation of sudden deaths among patients with seizures and epilepsy. Neurology. 2017;1–8.

6. Long L, Cotterman-Hart S, Shelby J. To reveal or conceal? Adult patient perspectives on SUDEP disclosure. Epilepsy Behav. 2018/07/14 ed. 2018 Sep;86:79–84. DOI: 10.1016/j.yebeh.2018.06.026.

7. Massey CA, Sowers LP, Dlouhy BJ, Richerson GB. Mechanisms of sudden unexpected death in epilepsy: the pathway to prevention. Nature Reviews Neurology. 2014;10:271–82. DOI: 10.1038/nrneurol.2014.64.

8. Ali A, Wu S, Issa NP, Rose S, Towle VL, Warnke P, et al. Association of sleep with sudden unexpected death in epilepsy. Epilepsy & Behavior. 2017;76:1–6. DOI: 10.1016/j.yebeh.2017.08.021.

9. Lamberts RJ, Thijs RD, Laffan A, Langan Y, Sander JW. Sudden unexpected death in epilepsy: people with nocturnal seizures may be at highest risk. Epilepsia. 2011/12/24 ed. 2012;53:253–7. DOI: 10.1111/j.1528-1167.2011.03360.x.

10. Ryvlin P, Nashef L, Lhatoo SD, Bateman LM, Bird J, Bleasel A, et al. Incidence and mechanisms of cardiorespiratory arrests in epilepsy monitoring units (MORTEMUS): a retrospective study. Lancet Neurol. 2013;12:966–77. DOI: 10.1016/S1474-4422(13)70214-X.

11. Alexandre V, Mercedes B, Valton L, Maillard L, Bartolomei F, Szurhaj W, et al. Risk factors of postictal generalized EEG suppression in generalized convulsive seizures. Neurology. 2015;85:1598–603. DOI: 10.1212/WNL.0000000000001949.

12. Liebenthal JA, Wu S, Rose S, Ebersole JS, Tao JX. Association of prone position with sudden unexpected death in epilepsy. Neurology. 2015;84:703–9.

13. Wang S, Graf S, Xu J, Issa NP, Ali A, Rose S, et al. The incidence of peri-ictal prone position in patients with generalized convulsive seizures. Epilepsy and Behavior. 2016;61:158–61. DOI: 10.1016/j.yebeh.2016.05.033.

14. Shmuely S, Surges R, Sander JW, Thijs RD. Prone sleeping and SUDEP risk: The dynamics of body positions in nonfatal convulsive seizures. Epilepsy Behav. 2016/08/05 ed. 2016 Sep;62:176–9. DOI: 10.1016/j.yebeh.2016.06.017.

15. Vilella L, Lacuey N, Hampson JP, Rani MRS, Sainju RK, Friedman D, et al. Postconvulsive central apnea as a biomarker for sudden unexpected death in epilepsy (SUDEP). Neurology. 2018/12/21 ed. 2019 Jan 15;92(3):e171–82. DOI: 10.1212/WNL.0000000000006785.

16. Vilella L, Lacuey N, Hampson JP, Zhu L, Omidi S, Ochoa-Urrea M, et al. Association of Peri-ictal Brainstem Posturing With Seizure Severity and Breathing Compromise in Patients With Generalized Convulsive Seizures. Neurology. 2020/12/04 ed. 2021 Jan 19;96(3):e352–65. DOI: 10.1212/WNL.0000000000011274.

17. Myers KA, Bello-Espinosa LE, Symonds JD, Zuberi SM, Clegg R, Sadleir LG, et al. Heart rate variability in epilepsy: A potential biomarker of sudden unexpected death in epilepsy risk. Epilepsia. 2018;59(7):1372–80. DOI: 10.1111/epi.14438.

18. Nayak CS, Sinha S, Nagappa M, Thennarasu K, Taly AB. Lack of heart rate variability during sleep-related apnea in patients with temporal lobe epilepsy (TLE)—an indirect marker of SUDEP? Sleep and Breathing. 2017;21:163–72. DOI: 10.1007/s11325-016-1453-6.

19. Thijs RD, Ryvlin P, Surges R. Autonomic manifestations of epilepsy: emerging pathways to sudden death? Nat Rev Neurol. 2021/10/31 ed. 2021 Dec;17(12):774–88. DOI: 10.1038/s41582-021-00574-w.

20. Galland B, Reeves G, Taylor B, Bolton D. Sleep position, autonomic function, and arousal. Arch Dis Child Fetal Neonatal Ed. 1998 May;78(3):F189–94.

21. Kato I, Scaillet S, Groswasser J, Montemitro E, Togari H, Lin JS, et al. Spontaneous Arousability in Prone and Supine Position in Healthy Infants. Sleep. 2006 Jun;29(6):785–90. DOI: 10.1093/sleep/29.6.785.

22. Richardson HL, Walker AM, Horne RSC. Sleep position alters arousal processes maximally at the high-risk age for sudden infant death syndrome. Journal of Sleep Research. 2008;17(4):450–7. DOI: 10.1111/j.1365-2869.2008.00683.x.

23. Esmaeili B, Dworetzky BA, Glynn RJ, Lee JW. The probability of sudden unexpected death in epilepsy given postictal prone position. Epilepsy Behav. 2021/02/12 ed. 2021 Mar;116:107775. DOI: 10.1016/j.yebeh.2021.107775.

24. Sowers LP, Massey CA, Gehlbach BK, Granner MA, Richerson GB. Sudden unexpected death in epilepsy: fatal post-ictal respiratory and arousal mechanisms. Respir Physiol Neurobiol. 2013/05/28 ed. 2013 Nov 1;189(2):315–23. DOI: 10.1016/j.resp.2013.05.010.

25. Richerson GB, Buchanan GF. The serotonin axis: Shared mechanisms in seizures, depression, and SUDEP. Epilepsia. 2011;52:28–38. DOI: 10.1111/j.1528-1167.2010.02908.x.

26. Guntheroth WG, Spiers PS. Sleeping prone and the risk of sudden infant death syndrome. JAMA. 1992/05/06 ed. 1992 May 6;267(17):2359–62.

27. Ponsonby AL, Dwyer T, Gibbons LE, Cochrane JA, Wang YG. Factors potentiating the risk of sudden infant death syndrome associated with the prone position. N Engl J Med. 1993/08/05 ed. 1993 Aug 5;329(6):377–82. DOI: 10.1056/NEJM199308053290601.

28. Gemble A, Hubert C, Borsa-Dorion A, Dessaint C, Albuisson E, Hascoet JM. Knowledge assessment of sudden infant death syndrome risk factors in expectant mothers: A prospective monocentric descriptive study. Arch Pediatr. 2019/12/01 ed. 2020 Jan;27(1):33–8. DOI: 10.1016/j.arcped.2019.10.012.

29. Changing concepts of sudden infant death syndrome: implications for infant sleeping environment and sleep position. American Academy of Pediatrics. Task Force on Infant Sleep Position and Sudden Infant Death Syndrome. Pediatrics. 2000/03/04 ed. 2000 Mar;105(3 Pt 1):650–6. DOI : 10.1542/peds.105.3.650.

30. Mitchell EA, Brunt JM, Everard C. Reduction in mortality from sudden infant death syndrome in New Zealand: 1986-92. Arch Dis Child. 1994/04/01 ed. 1994 Apr;70(4):291–4. DOI: 10.1136/adc.70.4.291.

31. Tao JX, Qian S, Baldwin M, Chen XJ, Rose S, Ebersole SH, et al. SUDEP, suspected positional airway obstruction, and hypoventilation in postictal coma. Epilepsia. 2010;51:2344–7. DOI: 10.1111/j.1528-1167.2010.02719.x.

32. McLean BN, Wimalaratna S. Sudden death in epilepsy recorded in ambulatory EEG. J Neurol Neurosurg Psychiatry. 2007/03/29 ed. 2007;78:1395–7. DOI: 10.1136/jnnp.2006.088492.

33. SJ P, M WY, VP S. Sudden death in epilepsy: single case report with video-EEG documentation. Epilepsia. 1992;33(S3):123.

34. Langan Y, Nashef L, Sander JW. Case-control study of SUDEP. Neurology. 2005;64:1131–3. DOI: 10.1212/01.WNL.0000156352.61328.CB.

35. Nashef L, Fish DR, Garner S, Sander JWAS, Shorvon SD. Sudden Death in Epilepsy: A Study of Incidence in a Young Cohort with Epilepsy and Learning Difficulty. Epilepsia. 1995;36:1187–94. DOI: 10.1111/j.1528-1157.1995.tb01061.x.

36. van Andel J, Thijs RD, de Weerd A, Arends J, Leijten F. Non-EEG based ambulatory seizure detection designed for home use: What is available and how will it influence epilepsy care? Epilepsy and Behavior. 2016;57:82–9. DOI: 10.1016/j.yebeh.2016.01.003.

37. Sveinsson O, Andersson T, Mattsson P, Carlsson S, Tomson T. Clinical risk factors in SUDEP: A nationwide population-based case-control study. Neurology [Internet]. 2019/12/14 ed. 2019 Dec 12; Available from: https://www.ncbi.nlm.nih.gov/pubmed/31831600DOI: 10.1212/WNL.0000000000008741.

38. Picard RW, Migliorini M, Caborni C, Onorati F, Regalia G, Friedman D, et al. Wrist sensor reveals sympathetic hyperactivity and hypoventilation before probable SUDEP. Neurology. 2017;10.1212/WNL.0000000000004208. DOI: 10.1212/WNL.0000000000004208.

39. Catcheside PG, Mohtar AA, Reynolds KJ. Airflow resistance and CO2 rebreathing properties of anti-asphyxia pillows designed for epilepsy. Seizure. 2014;23:462–7. DOI: 10.1016/j.seizure.2014.03.007.

40. Kinney HC, Thach BT. The sudden infant death syndrome. N Engl J Med. 2009/08/21 ed. 2009 Aug 20;361(8):795–805. DOI: 10.1056/NEJMra0803836.

